# Modified SIR-model applied to covid-19, similarity solutions and projections to further development

**DOI:** 10.1101/2020.07.30.20165035

**Authors:** E. Rebhan

## Abstract

The SIR-model is adapted to the covid-19 pandemic through a modification that consists in making the basic reproduction number variable. Independent of it, another reproduction number is introduced, which is defined similarly to the usual net reproduction number. Due to its simple analytic form, it enables a clear interpretation for all values. A further parameter, provisionally called acceleration parameter, is introduced and applied, which enables a more differentiated characterization of the infection number dynamics. By a variable transformation the 3 equations of the modified SIR-model can be reduced to 2. The latter are solved up to ordinary integrations. The solutions are evaluated for current situations, yielding a pretty good match with the data reported. Encouraged by this, a variety of possible future developments is examined, including linear and exponential growth of the infection numbers as well as sub- and super-exponential growth. In particular, the behavior of the two reproduction numbers and the acceleration parameter is studied, which in some cases leads to surprising results. With regard to the number of unreported infections it is shown, that from the solution for a special one solutions for others can be derived by similarity transformations.

## 1 Introduction

In this paper it is shown that the SIR-model [1] can pretty well be adjusted to the data reported for the covid-19 pandemic by means of a modification, which consists in allowing a dynamic variability of the usually constant basis reproduction number *R*_0_. An additional reproduction number is introduced which is similarly defined as the sensitive R-number of the Robert Koch Institute (RKI) or the net reproduction rate. Another number turns out to be particularly useful, the values of which allow even more differentiated informations about the progression of the infection numbers. For it, the term acceleration parameter is proposed.

First, by a suitable transformation of variables the system of 3 coupled nonlinear differential equations of the SIR-model is converted into a system with only 2 equations. The determination of the general solution of the latter is then carried on so far that only ordinary integrations remain. This makes it unnecessary to develop a numerical program for the solutions. Rather, the simplicity and clarity of the solutions obtained make it possible to quickly achieve results for concrete problems, especially when programs like MATHEMATICA are employed. With regard to the number of unreported infections it turns out, that despite the non-linearity of the equations, from the solution for a special one the solutions for other numbers can be derived by similarity transformations under rather unobtrusive assumptions.

In applying the solutions to the covid-19 pandemic, the reported data for one variable are replaced by a best least-squares fit function and thus integrated in the expanded SIR-model. Thereupon, from this the associated solution for a further variable is determined and compared with the corresponding reported data. It turns out that at least for more advanced states there is fairly good agreement. This encourages solutions to be explored, that continue preceding solutions to some extent into the future. Various assumptions about the further course of the infection or reproduction numbers are thus examined and compared. Also considered are the conditions under which the pandemic comes to an end. In the concluding section, some more qualitative aspects of the pandemic are discussed.

In order to keep the health damage and death cases caused by the pandemic as low as possible, significant restrictions and unfamiliar actions are expected of the population. So that people understand why these are necessary, it is important that the reasons for them are communicated as precise and intelligible as possible. This in turn presupposes that the knowledge about the various aspects of the pandemic is as extensive as possible, while being precise and thorough. This article was written with the intention and hope of making a small contribution to this. It is mainly based on the system of SIR-equations which were taken from Wikipedia [2]. Therefore, only a few references are given.

## 2 Definitions, basic equations and solutions

### 2.1 Definitions and basic equations

Neglecting birth and non-covid death rates, the basic equations of the SIR-model are

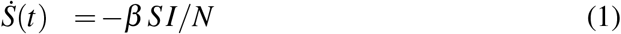

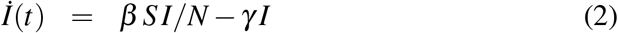

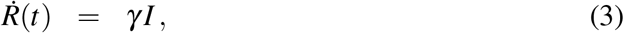

where the following notations are used: *S*=susceptible individuals, *I*=infectious individuals, *R*=removed individuals (recovered with acquired immunity to the disease or deceased), *N*=(invariable) total number of individuals, *t*=time with the unit day, and 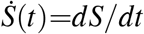 etc. Furthermore, *γ* is the daily recovery rate of infected individuals, and *β* is the daily rate of new infections caused by an infected individual. The SIR-model is based on the assumptions that individuals can be infected only once, become contagious immediately after infection and remain so until they gain immunity or die. The time-dependent variables of the SIR-equations satisfy the relation

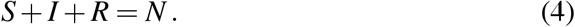

We also use the basic reproduction number

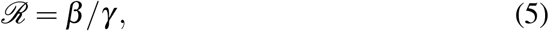

thereby omitting the subscript 0 of the usual notation *R*_0_ in view of a modification specified further on.

Clearly, the numerical solution of the above equations is not a problem. However, considerable simplifications are possible that make it easier to answer specific questions and enable a better understanding of solution properties. In a first step, the system of 3 equations (1)-(3) can be reduced to a system of 2 equations by introducing the variable

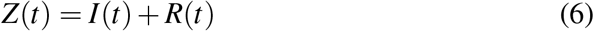

which is also used in the infection diagrams of institutions determining the infection numbers like the Robert Koch Institute (RKI) or the Johns Hopkins University (JHU). Eliminating *S* and *R* by use of *S*=*N* −*Z* and *R*=*Z*− *I* (which follows from Eqs. (4) and (6)), and using *β* =*γℛ* (which follows from Eq. (5)), Eqs. (2)-(3) become

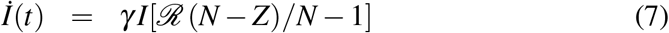

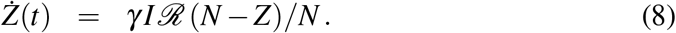

With the definitions

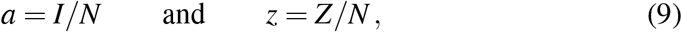

we switch from the extensive variables *I* and *Z* to the intensive variables *a* and *z*, thus finally ending up with the equations

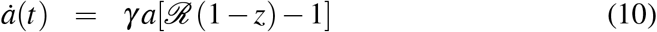

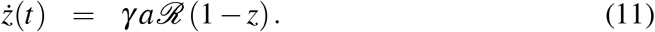

With the help of Eqs. (4), (6) and (9), the solutions *a*(*t*) and *z*(*t*) of these equations immediately lead to *S*(*t*)=*N*(1−*z*(*t*)) and *R*(*t*)=*N*(*z*(*t*)−*a*(*t*)).

### 2.2 Variable basic reproduction number and solutions

Dividing Eq. (10) by Eq. (11), with 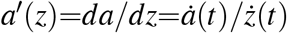 we obtain

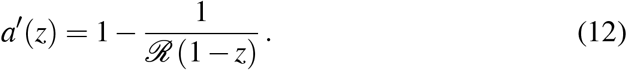

For fixed *ℛ* this is a differential equation for *a* with the solution

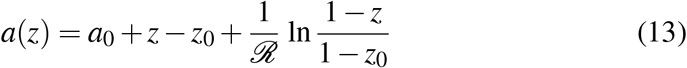

to the initial condition *a*(*z*_0_)=*a*_0_.

We also want to consider situations in which *a*(*t*) and/or *z*(*t*) are predetermined by empirical data on the progress of the pandemic. In this case Eq. (13) cannot be expected to be a suitable solution. However, it turns out that the SIR-model can be adapted to the covid-19 pandemic by allowing the basic reproduction number to be variable. In practice, such changes are brought about by ordering measures to protect the population, such as quarantine, social distancing, hygienic washing of the hands or mandatory wearing of respirators, but also through sensible and responsible behavior of the population. Only experience can teach how this quantitatively affects *ℛ* or the reproduction number *R*_*z*_ introduced further down. Specifically, we introduce the replacement *ℛ* → *ℛ*(*z*), where *ℛ*(*z*) is obtained by resolving Eq. (12) with respect to *ℛ*,

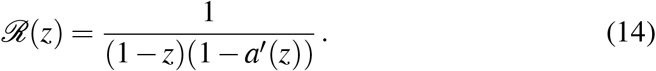

Inserting this in Eq. (11) yields

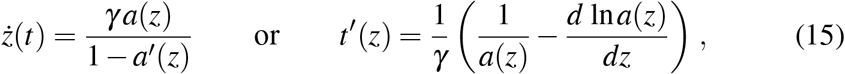

where *t*(*z*) is the inverse function of *z*(*t*). *ż*(*t*) is the temporal increase of *z*(*t*), which can be looked up in the daily records of the RKI [3] or JHU [4]. Integration of Eq. (15b) ^1^ yields

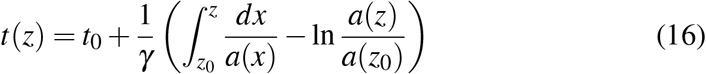

with *t*(*z*)=*t*_0_ for *z*=*z*_0_. (Eqs. (15)-(16) also apply to the case of fixed *ℛ*.)

Eq. (16) is the solution for given *a*(*z*). We also consider the case where *t*(*z*) is given and a solution for *a*(*z*) is sought. The differential equation from which *a*(*z*) must be determined is Eq. (15a), which with *ż*(*t*)=1*/t*^*′*^(*z*) can be converted into

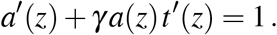

The solution to this equation is readily obtained by using the method of variation of constants and is

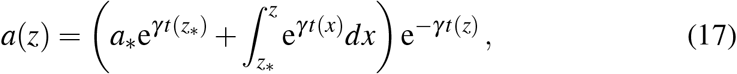

where *a*_∗_ and *z*_∗_ are arbitrary fixed values in the range of possible *a*- and *z*-values.

**Annotation:** From a mathematical point of view, the modification *ℛ*= const to *ℛ*=*ℛ*(*z*) in the SIR-model means that the system (10)-(11) of 2 equations is under-determined because it contains 3 unknown functions, *z*(*t*), *a*(*t*) and *ℛ*(*z*). However, this is not a problem because it would not even be desirable for *ℛ*(*z*) to be determined by the equations. In practical applications, there is the possibility of specifying any of the 3 functions, either by adaption to empirical data or by making assumptions about the future development

### 2.3 Additional reproduction number

The variable reduction number introduced in Eq. (14) does not allow a clear interpretation of its values, even though we will see later on that it has values and time variations similar to usual ones. For better comparison with the latter and to enable better interpretability, we introduce a second reproduction number that is similarly defined as the sensitive R-number of the RKI or the net reproduction rate.

For this purpose we assume that a list of successive daily values *Z*_*n*_ for the total number of infected individuals is given. First, a best least-squares fit function *z*(*t*) [5] to the corresponding data {*t*_*n*_, *z*_*n*_}, *n*=0, 1, 2, … [6] is constructed, which in many cases can be done using a power series 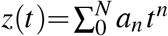. (In this way, the data is smoothed, as is also done in a similar way in the usual calculation of R values by averaging). The current increase in *z*(*t*) is *ż*(*t*) Δ*t*, and furthermore

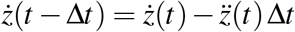

holds. With this, we define our new reproduction factor as

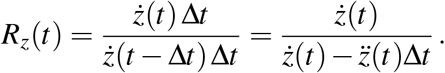

We still relate it to the time unit *day* by putting Δ*t*=1, and after a slight reshaping we finally get

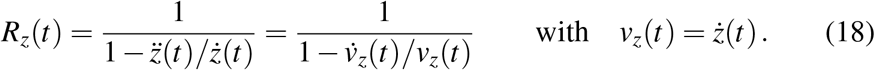

(*v*_*z*_ is the growth velocity of *z*(*t*).) On cursory inspection, it looks like *R*_*z*_(*t*) is an instantaneous value related only to the moment *t*. Contrary to this, in the calculation of the sensitive *R*-value of the RKI, due to a time delay in the reported infection data past values are included, and the data are smoothed by averaging over several days to compensate for statistical fluctuations. On closer inspection, however, this also applies to our *R*_*z*_(*t*) because to determine the function *z*(*t*) via a best least-squares fit to a data set {*t*_*n*_, *z*_*n*_}, past data are included as well as smoothing takes place.

For the *R*-number of RKI and the usual net *R*-number the value 1 is easy to understand, because on average every infected individual infects another one during its contagious stage, whence the number of newly infected individuals remains constant. For values ≠ 1 a similarly simple explanation is not possible, except that a deviation downwards or upwards is more favorable or unfavorable the larger it is. For our *R*_*z*_-number the situation is slightly better, because Eq. (18) can be used to quantify the conditions under which a deviation increases or decreases: the greater the relative acceleration 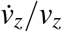, the greater the deviation from 1 where *R*_*z*_(*t*)≡1 for constant *ż*(*t*) as above. It would even be possible to use the number

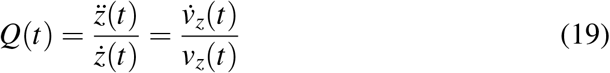

as another number for characterizing the state of the pandemic. However, in the next section it is shown that a modification of *Q*(*t*) is a much better choice.

We would like to apply our reproduction number also where not *z*(*t*) is specified but its inverse *t*(*z*). For this purpose, we differentiate the identity t′(*z*) *ż*(*t*) = (*dt/dz*)*/*(*dz/dt*) ≡ 1 with respect to *t*, obtaining 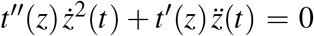 or 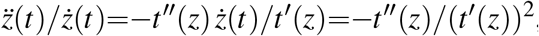, and by insertion into Eq. (18) get

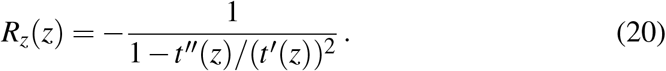

For the case of given *a*(*z*) addressed in Section 3.1.1, we still determine how *ℛ*(*z*) can be expressed in terms of *a*(*z*). From Eq. (11) with *ℛ → ℛ*(*z*) we get

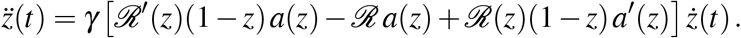

Dividing by *ż*(*t*) and using

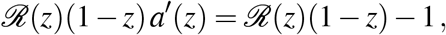

which follows from Eq. (14), we get

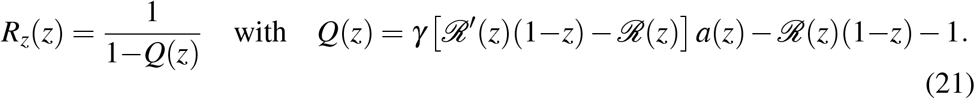

*γ* must be determined so that *R*_*z*_(*z*_0_) coincides with the *R*_*z*_(*t*_0_) of Eq. (18).

### 2.4 Acceleration parameter

The terms linear and exponential growth are often used to characterize the increase in the number of infections, the net reproduction number being used to distinguish them. As we will see, this is a rather vague and possibly misleading characterization. In the following, another parameter is proposed that enables a more differentiated assessment.

Combining Eq. (18) and Eq. (19) yields

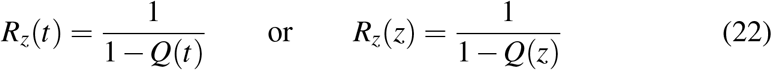

from which follows that *R*_*z*_ and *Q* offer the same information content. The acceleration parameter proposed by us consists in a modification of 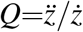 and is

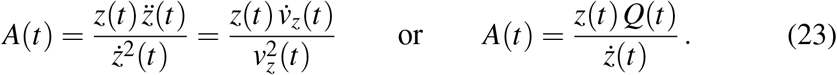

The relationship between *A*(*t*) and *Q*(*t*) shows that *A*(*t*) contains more information than *Q*(*t*).

Now, we first calculate *A* (*t*) for the case of exponential growth, *z*(*t*)=*z*_0_e^*α t*^. With *ż*(*t*)=*α z*(*t*) and 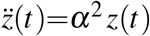 we get *A*(*t*) ≡1. For other growth patterns we expand *z*(*t*) around an arbitrary *z*_0_=*z*(*t*_0_) according to

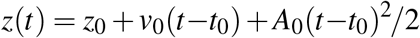

and get with *ż*(*t*)=*v*_0_+*A*_0_ (*t*−*t*_0_) and 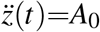 for *t* → *t*_0_ the value

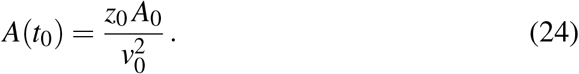

Let us now consider different values of *A*_0_ for the same values of *z*_0_ and *v*_0_. In the case of exponential growth we have *v*_0_ = *αz*_0_, *A*_0_ = *α*^2^ *z*_0_ and from this 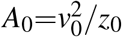 since *A*(*t*_0_)=1. For 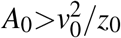(i.e. *A*(*t*_0_)<1, the curve *z*(*t*) is curved stronger upward than an exponential curve of the same slope through the same point *z*_0_ and therefore (locally) shows a super-exponential growth. For 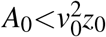, it is less curved and shows a sub-exponential growth. For *A*_0_=0 it is not curved and (locally) linear, and for *A*_0_*<*0 its growth is slower than linear and stops growing at all for *v*_0_→ 0 with the consequence *A*(*t*_0_) → − ∞. These results are summarized in the formula

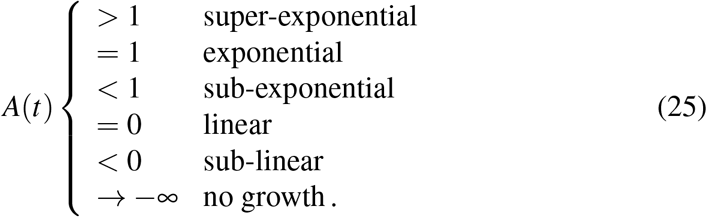

Applications are presented in later sections.

We still want to find out how *A*(*t*) and *R*_*z*_(*t*) are related. According to Eq. (23b) we have *A*=*Qz/ż*, and from Eq. (22) follows *Q*=(*R*_*z*_ −1)*/R*_*z*_. Combining these results yields

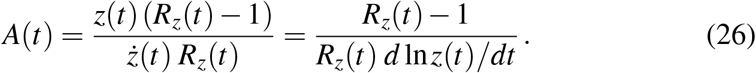

The function *z*(*t*), required to calculate *A*(*t*), is determined as discussed in the last section. If, as in the practical determination of the net reproduction number, only the data from the last 8 days are used, a best fit in the class of polynomials 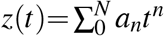 should provide suitable results. Daily updating is easy to do by omitting the oldest from the data and adding the newest. Although the parameter *A*(*t*) provides more information than the reproduction number *R*_*z*_(*t*), it is also not sufficient for a complete assessment of the stage of the pandemic. For decisions on measures to its containment or about the relaxation of such measures, still other quantities (like *ż*(*t*) etc.) and results of statistical investigations must be consulted.

### 2.5 Various growth possibilities of *z(t)*

#### 2.5.1 Linear growth

The general solution for linear growth is

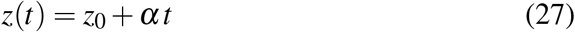

with constant *z*_0_ and *α*. Inserting the resulting relations *ż*(*t*)=*α* and 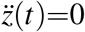 in Eq. (18) yields

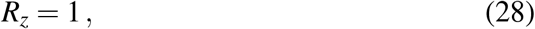

and vice versa Eq. (27) follows from Eq. (28).

Inserting *ż*(*t*)=*α* in Eq. (15a) leads to *α*=*γ a*(*z*)*/*(1−*a*′(*z*)) or

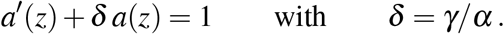

The solution to this equation is *a*(*z*)=1*/δ* so that *a*′ (*z*)=0, and with this Eq. (14) delivers

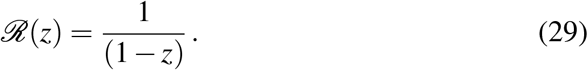

*R*_*z*_ and *ℛ* agree only for *z*=0. In order to keep the number of infectious individuals (*I* or *a* resp.) constant, *R*_*z*_ must be held fixed at the value 1, while *ℛ* increases continuously according to Eq. (29) with Eq. (27).

#### 2.5.2 Exponential growth

The general solution for exponential growth is

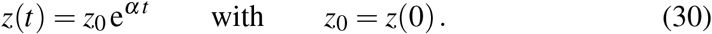

with With *ż*(*t*)=*α z*(*t*) and 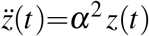 Eq. (18) yields

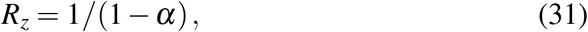

and from Eq. (11) we get

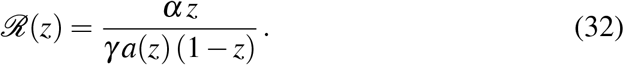

In the last result *a*(*z*) still has to be calculated what Eq. (17) can be used for. The inverse function *t*(*z*) to *z*(*t*) from Eq. (30) required for this is

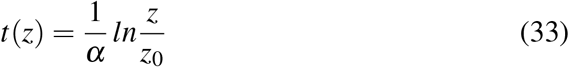

so that

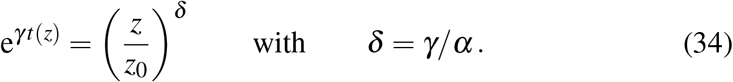

According to this *t*(*z*_0_)=0. Choosing *z*_∗_=*z*_0_ and *a*_∗_=*a*_0_ in Eq. (17), inserting in it the result (32), and using

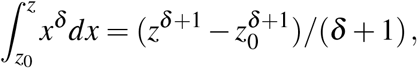

we finally get

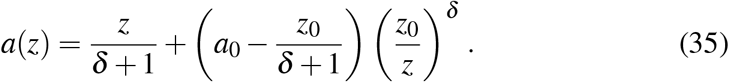

When this is inserted in Eq. (32), the solution *ℛ*(*z*) is complete. The time behavior of *ℛ* is obtained by substituting *z*(*t*) from Eq. (30) for *z* in *ℛ*(*z*).

#### 2.5.3 Accelerated growth

We are also interested in how constant acceleration or deceleration 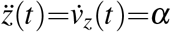 of *z*(*t*) affects the reproduction number *R*_*z*_. In this case we have

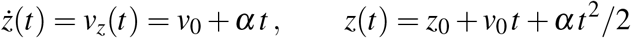

And

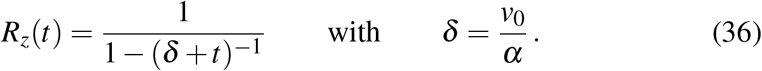

For *α>*0 we have *δ>*0, *R*_*z*_(0)=1*/*(1− 1*/δ*)*>*1 and *R*_*z*_(0)*>R*_*z*_(*t*)*>*1. Asymptotically we get

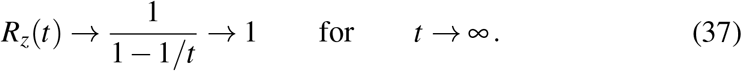

In spite of an accelerated increase of *z*(*t*) the reproduction number *R*_*z*_ decreases. This illustrates that the specification of a reproduction number, *R*_*z*_ in our case, is not sufficient to characterize the pandemic.

For *α<*0 or *δ<*0 resp. we can write

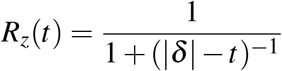

whence *R*_*z*_(*t*)*<R*_*z*_(0)*<*1, and asymptotically we get

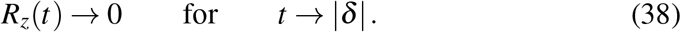

#### 2.5.4 Growth with fixed *A(t)*

For fixed *A*(*t*)=const=*A*, a conversion of Eq. (23) with use of *v*_*z*_=*ż*(*t*) yields

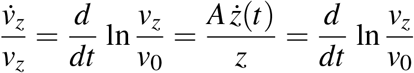

and after integration

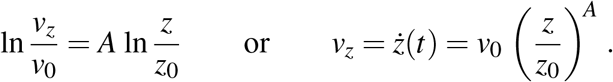

Further integration leads to the result

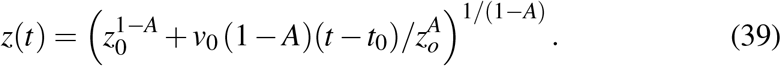

Fig. 1 shows *z*(*t*) for 5 different values of *A* as an illustration, the initial values *z*_0_ and *v*_0_ being arbitrarily chosen but equal for all curves *z*(*t*).

**Figure 1:**
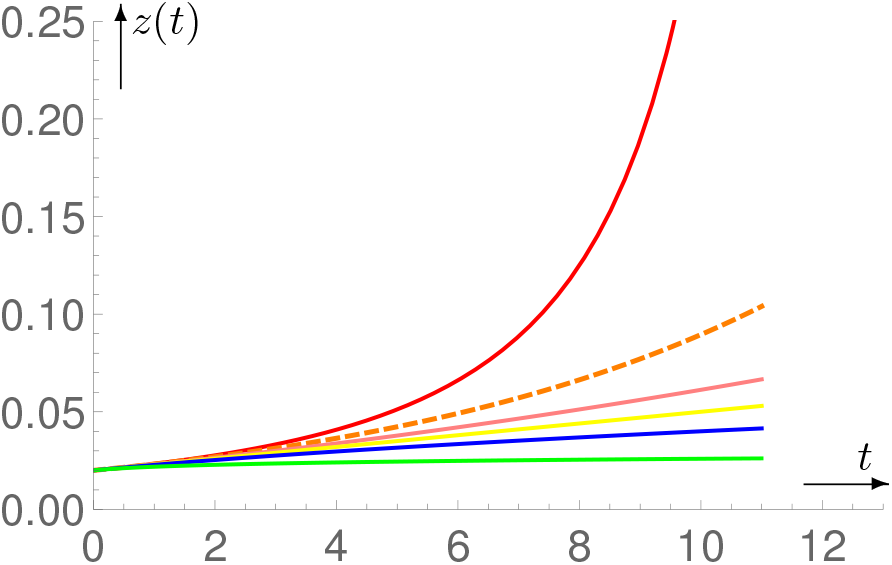
*z*(*t*) for *A* = 1.5, 1, 0.5, 0, − 10 in the order from top to bottom, the curve for exponential growth being dashed.

**Annotation:** Linear, exponential and super-exponential growth are all possible, but due to *z*(*t*) *≤* 1 they can only occur temporarily.

### 2.6 Linear growth of *R_z_ (t)*

After the surprising result that *R*_*z*_(*t*) decreases when *z*(*t*) is accelerated, we are interested under which circumstances *R*_*z*_ increases, and examine which consequences result from the ansatz

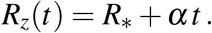

With this we get from Eq. (18)

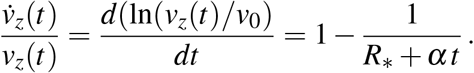

Integration with respect to t results in

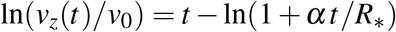

Or

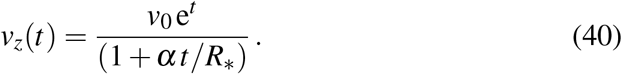

*v*_*z*_(*t*) growth faster than exponentially (super-exponential growth) for increasing *R*_*z*_(*t*) (positive *α*) and slower than exponentially (sub-exponential growth) for decreasing *R*_*z*_(*t*) (negativ *α*).

### 2.7 Similarity solutions

The data published by the RKI or JHU and updated daily refer to the reported cases. Since many individuals are infected and do not know because they have no symptoms of illness, there is a high number of unreported cases. This can be taken into account by multiplying the reported data by factors. In the equations of the modified SIR-model then the substitutions 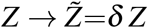 with constant *δ* must be made. Eq. (6) must be satisfied by both sets of variables, which means that

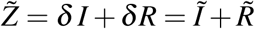

must apply. We now make the (not compulsory) assumption that *Ĩ*=*δ I* holds, from which 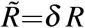 *R*follows. (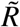 is the sum of those who have recovered and those who have died. Since the latter are generally well known, practically none of them are unreported. For the validity of 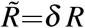, this would have to be compensated by multiplying the recovered ones by a somewhat higher factor.) To the specific variables *a* and *z* the same relationships apply as for *I* and *Z*, i.e.

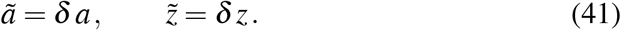

The multiplying factor *δ* is not known, but can be estimated ^2^ and be quite high (e.g. 10 or even higher). It would be an unpleasant surprise if, after completion of a calculation, it turned out that the factor *δ* used for it was wrong and the whole calculation had to be repeated with another *δ*. The following shows that this can be avoided under the above conditions.

Let *a*(*z*) be the function resulting from a concrete reported data set {*a*_*n*_, *z*_*n*_} via a best least-squares fit. The unreported cases are taken into account through the replacements *a_n_* → *ã*_*n*_ = *δ a_n_*, and 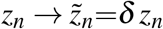 and for the associated fit function applies 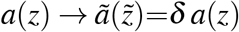 with 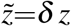 in short

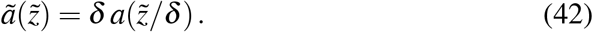

(From this follows 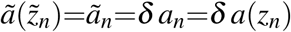 as required.) Using 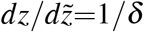, the derivation of this with respect to 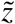 yields

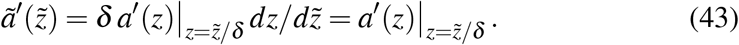

Eq. (14) must also apply to *ã* and 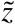, why with use of Eqs. (41) and (43) we get

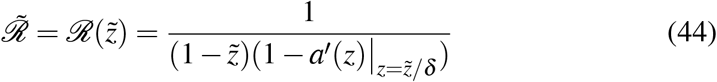

And

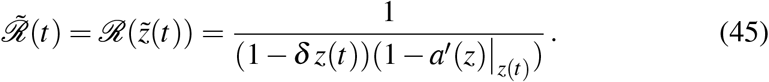

With 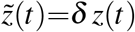, from Eqs. (18) and (19) we obtain

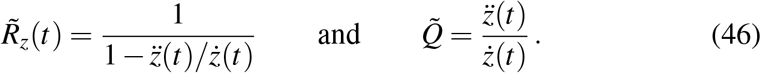

Finally we want to find out how the result (16) is influenced by *δ*. From Eq. (42) follows

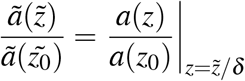

and

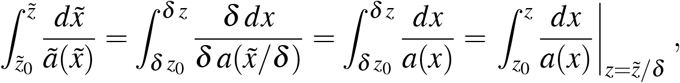

which eventually leads to the result

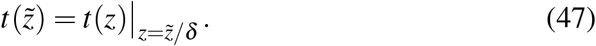

## 3 Investigation of concrete situations

### 3.1 Modified SIR-model adapted to specific cases

The SIR-model describes the average behavior of the statistical variables *S, I* and *R* or *z* and *a* in our case. Good agreement with the data supplied by the RKI or JHU can therefore only be expected when the number of cases is sufficiently high. Our approach to checking the applicability of the modified SIR-model consists in either specifying the function *a*(*z*) and calculating the function *z*(*t*) or *t*(*z*) resp. from the SIR-equations, or vice versa. To be more precise, we first look for an analytic function *a*(*z*) (or *t*(*z*)), whose values *a*(*z*_*i*_) (or *t*(*z*_*i*_)) at the positions *z*_*i*_ agree as well as possible with the given data pairs {*a*_*i*_, *z*_*i*_}=, *i*=0, 1, 2, … (or {*t*_*i*_, *z*_*i*_}, *i*=0, 1, 2, …). The function *t*(*z*) calculated from Eq. (16) (or *a*(*z*) calculated from Eq. (17)) is then compared with the corresponding reported data. The parameter *γ*, which has not yet been determined, is used as a fit parameter.

There are several problems with the described procedure regarding accuracy. In our calculations, we arbitrarily determined the number of unreported cases so that all reported data *a*_*i*_ and *z*_*i*_ are multiplied by the same factor *δ* = 10. As more and more people get tested for Covid-19, that number gets smaller, which we didn’t take into account. Furthermore, the data *a*_*i*_ of the infectious individuals are less reliable than the total number *z*_*i*_ of infections, which may be the cause why they are not specified in the official dashboards (see e.g. [3] or [4]). Alos, the determination of the functions *a*(*z*) or *z*(*t*) adapted to the given data is not straight forward. The path we have chosen avoids too much effort and may not be the best. However, in view of the other error sources, it is not worth investing too much effort into it.

#### 3.1.1 Given *a(z)*

We examine the case of given data for the function *a*(*z*), using the data reported for the world as an example and multiplying them by *δ* =10 to account for unreported cases. Corresponding data for the time interval from 3.1.2020 to 7.9.2020 (128 days) are taken from Ref. [7], and the function *a*(*z*) is the best least-squares fit in the class of polynomials 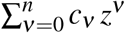, both plotted in Fig. 2.^3^ The function *a*(*z*) was then used to calculate *t*(*z*) from Eq. (16), *γ* being determined such that *t*(*z*_*i*_)=*t*_*i*_ for a specific pair {*z*_*i*_, *t*_*i*_} of the data set from Ref. [7]. The comparison of the solution *t*(*z*) with this data is also shown in Fig. 2. The correspondence is not perfect, but good enough that one can infer the usefulness of the modified SIR-model for certain questions, at least in terms of quality. Furthermore, the reproduction number *ℛ*(*z*) is shown in Fig. 3. (*R*_*z*_ is shown as a function of *t* in Fig. 5.)

**Figure 2:**
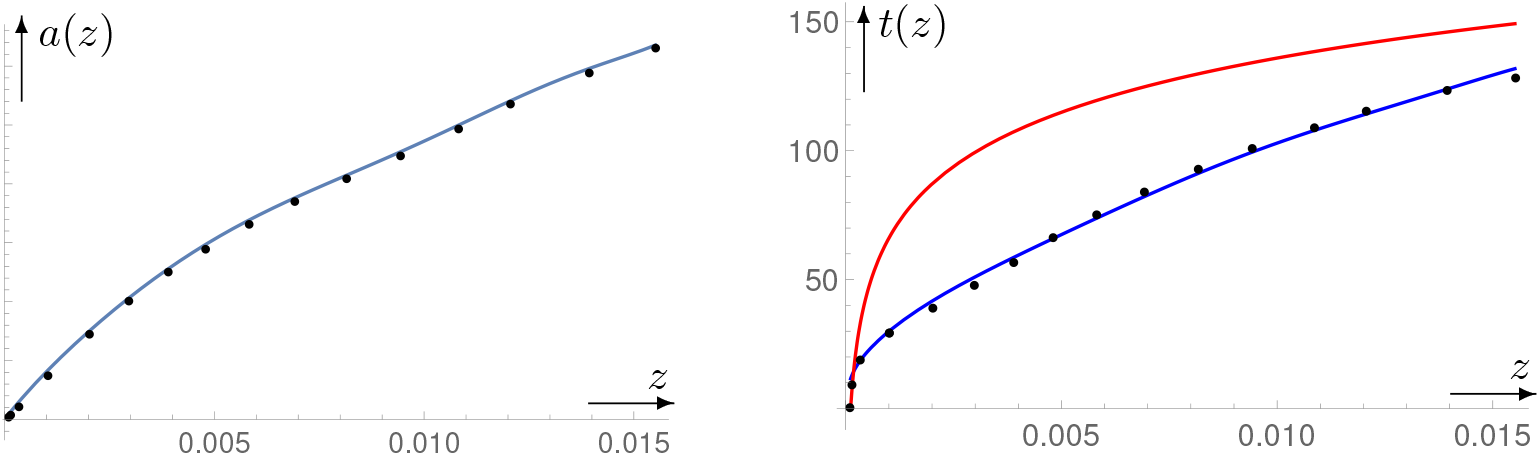
Left figure: reported data {*z*_*n*_, *a*_*n*_} (black dots) for the world and associated best fit curve *a*(*z*). Right figure: curve *t*(*z*) calculated from *a*(*z*) with Eq. (16) together with the corresponding reported data {*z*_*n*_, *t*_*n*_}. An exponential curve with the same initial values is entered above it, showing, that *z*(*t*) is initially super-exponential.

**Figure 3:**
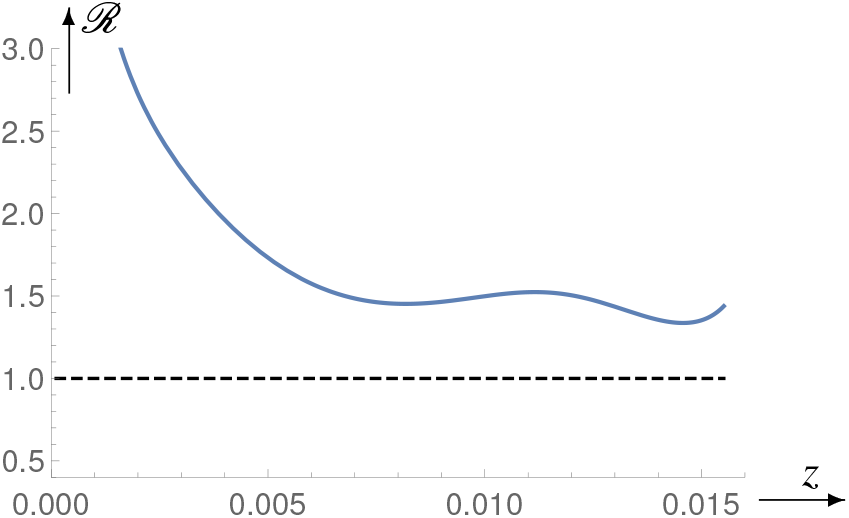
*ℛ*(*z*) and for the world, same time interval as in Fig. 2.

**Figure 4:**
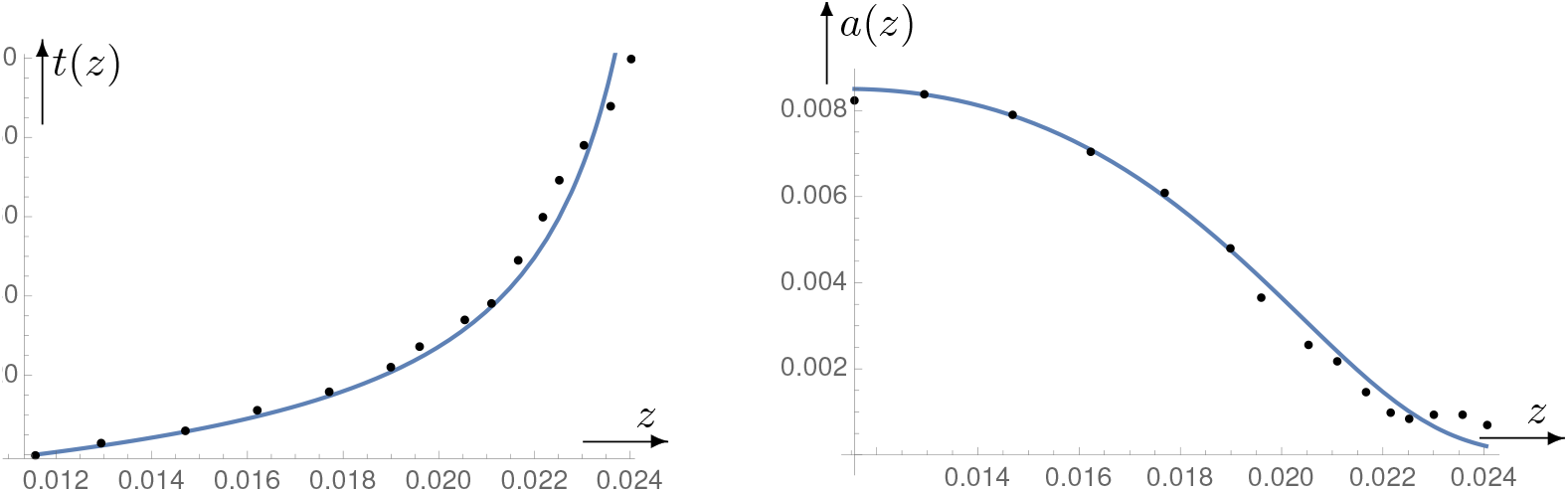
Left figure: reported data {*z*_*n*_, *t*_*n*_} for Germany and associated fit curve *t*(*z*). Right figure: curve *a*(*z*) calculated from *t*(*z*) with Eq. (17) together with the reported data {*z*_*n*_, *a*_*n*_}.

**Figure 5:**
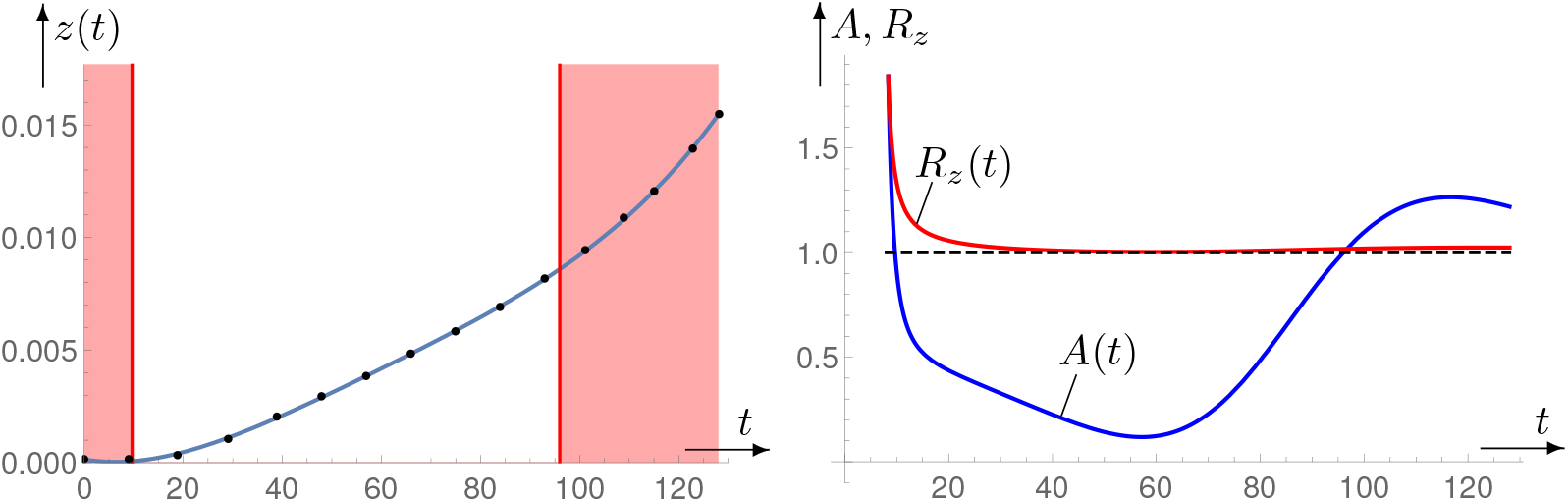
Left figure: reported data {*t*_*n*_, *z*_*n*_} for the world over a time interval of 128 days, and associated best fit curve *z*(*t*). In the shaded areas, *z*(*t*) is superexponential. Right figure: Associated parameters *A*(*t*) and *R*_*z*_(*t*) calculated from *z*(*t*) using Eqs. (18) and (23).

#### 3.1.2 Given *t(z)*

The reverse case, in which the data for *t*(*z*) are given and *a*(*z*) is calculated, is examined by the example of Germany. Corresponding data for the time interval from 4.4.2020 to 7.13.2020 are taken from Ref. [8]. Instead of *z*(*t*), we determine the inverse function *t*(*z*) as a fit to the reported data, but this time not in the class of polynomials, because this resulted in a function with too many fluctuations. (For similar reasons, only the range with decreasing values of the target function *a*(*z*) was examined.) Instead, the approach

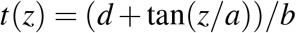

was used, the parameters *a, b*, and *d* being chosen to optimize the fit to the data (see Fig. 4). *a*(*z*) was then calculated from Eq. (17) and *γ* determined such that *a*(*z*_*i*_)=*a*_*i*_ for a specific pair {*z*_*i*_, *a*_*i*_} of the data. The comparison of the solution *a*(*z*) with the data is also shown in Fig. 4.

### 3.2 Application of the acceleration parameter *A(t)*

The acceleration parameter *A*(*t*) can be calculated directly from reported data without recourse to the SIR-model, the function *z*(*t*) again being determined by a (best) fit. Fig. 5 illustrates this process, using again the example of the world. For *A*(*t*)*>*1, the growth becomes super-exponential, and almost linear around day 60.

## 4 Future projections

We have seen that the modified SIR-model is well applicable to the pandemic, although in some cases rather only to its more advanced stage. We therefore assume that it can also be applied to future developments to a certain extent. Of particular interest is how *a*(*t*) and *z*(*t*) develop when certain assumptions about the further course of *ℛ*(*t*), *R*_*z*_(*t*) or *A*(*t*) are made.

### 4.1 Forecast solutions

**Predefined** *ℛ****(z)*:** In this case, from Eq. (10) results the equation

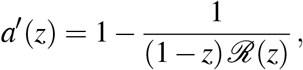

which is solved by

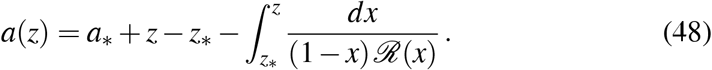

The associated solution *z*(*t*) is given by its inverse function *t*(*z*) to be calculated from Eq. (16). For *γ*, the value of the preceding solution can be used, to which the forecast solution is linked.

**Predefined *R***_*z*_***(t)*:** For given *R*_*z*_(*t*), from Eq. (18) results the equation

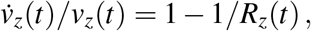

which is solved by

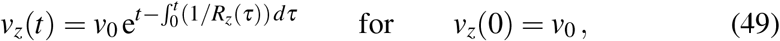

and integration of *ż*(*t*)=*v*_*z*_(*t*) yields

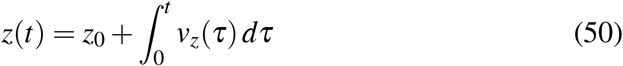

with *v*_*z*_(*τ*) given by Eq. (49). For determining the associated function *a*(*t*), we draw on Eq. (15a) in the form

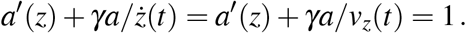

Using 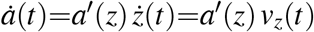 or 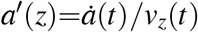, from this we get the equation

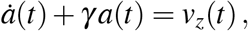

which is solved by

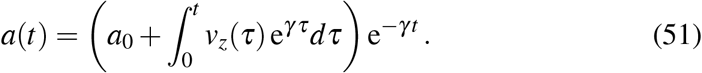

**Figure 6:**
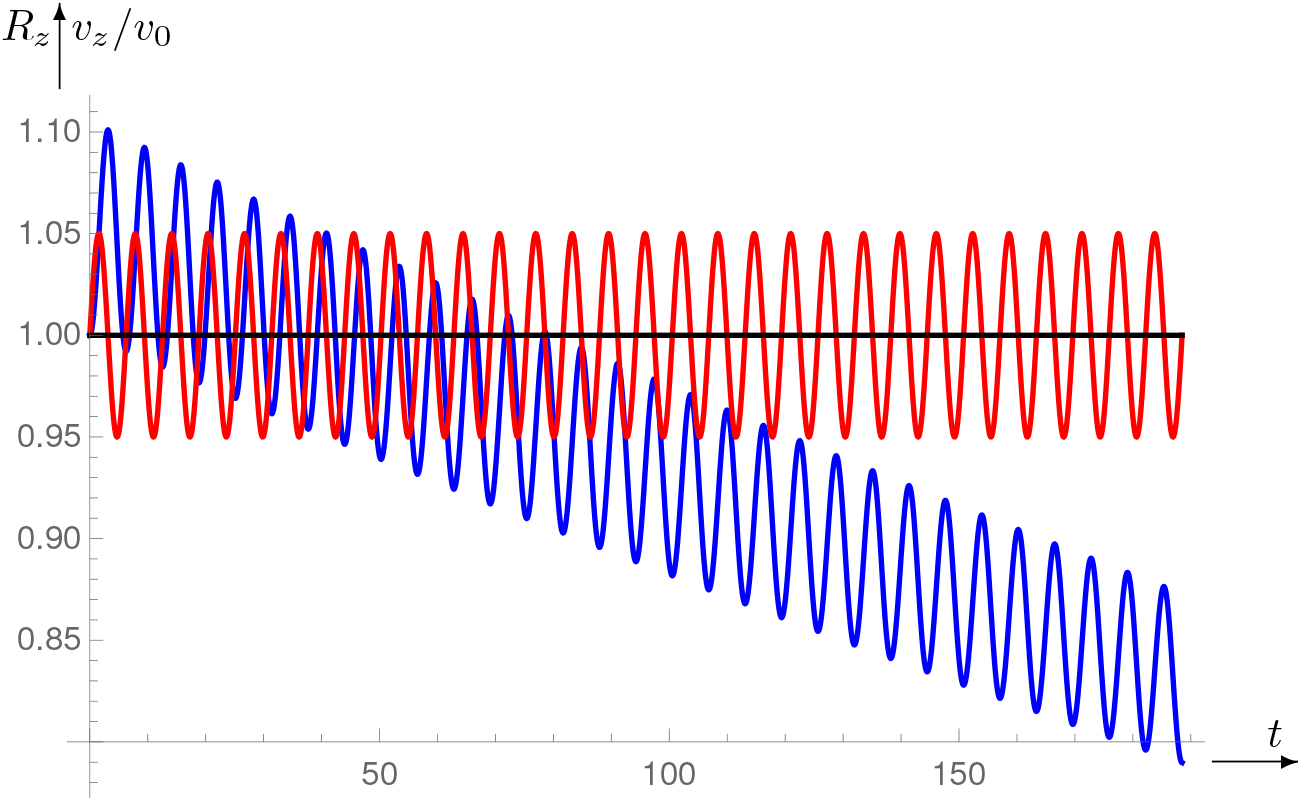
The horizontal curve is *R*_*z*_(*t*), the top-down curve *v*_*z*_(*t*)*/v*_0_.

***R***_*z*_**(t) oscillations around R**_*z*_**=1:** In practice, after the number of newly infected individuals has dropped to a lower level, it is variously observed that the net reproduction number is close to 1 and alternately exceeds this value and falls below it. We simulate this situation by making the ansatz

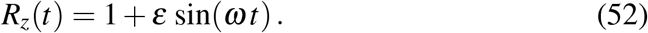

According to this and Eq. (49) the relative rate of new infections is

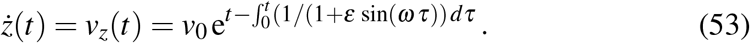

The integral in the exponent can indeed be expressed through analytic functions,

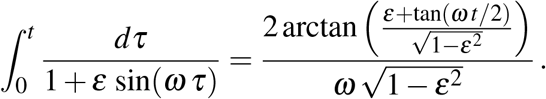

It is, however, rather tedious to get the various branches of the trigonometric functions aligned correctly adjacent to each other. We have therefore evaluated Eq. (53) numerically. The result is shown in Fig. 5. The surprising outcome is that *v*_*z*_(*t*) gradually decreases. The upward exceedings of limit 1 by *v*_*z*_ are therefore harmless if they are compensated for by undershoots of the same duration and strength.

### 4.2 End of the pandemic

A pandemic ends when there are no more infectious people, i.e. for *I*=0 or *a*=0.

**Case of constant** *ℛ*: In this case Eq. (13) applies, and for *a*=0 we obtain

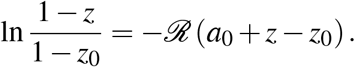

With the definitions

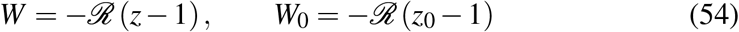

and by transition from the logarithm to the exponential function this results in

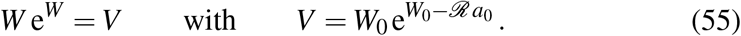

The resolution of this equation with respect to W is called Lambert-W-function, omega function or product logarithm. It has several branches, of which the one we need is called the principal solution *W*_0_(*V*). Thus, in terms of *W* and *V* the solution of our problem is *W* =*W*_0_(*V*). Returning to our original variables by use of Eqs. (54) and (55b), for the value *z*=*z*_*f*_, at which the pandemic ends, we get

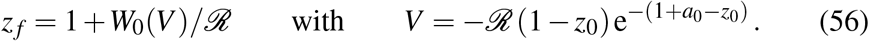

**Case of constant** *R*_*z*_: According to Eq. (18) for constant *R*_*z*_ we get

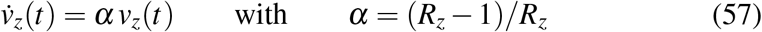

and from this

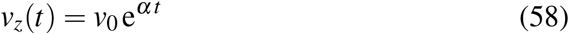

with *v*_0_ ≥ 0, because *z*(*t*) cannot decrease. This means that the constancy of *R*_*z*_ implies exponential growth of *v*_*z*_(*t*). According to Eq. (11) the condition *a*=0 for ending the pandemic calls for *ż*(*t*)=*v*_*z*_=0 which is not possible with *α* ≥ 0, and for *α<*0 this is only achieved after an infinitely long t ime. With *v* _*z*_=*ż*(*t*) from Eq. (58) results the differential equation *ż*(*t*)=*v*_0_ e^*α t*^ which is solved by

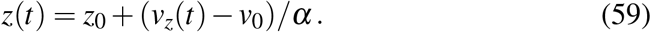

According to the termination condition *v*_*z*_=0, from this and Eq. (58) we obtain

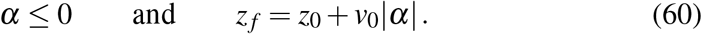

The final value *z* _*f*_ that *z*(*t*) assumes at the end of the pandemic remains below its maximum 1 only for |*α*|*>v*_0_*/*(1 −*z*_0_).

## 5 Concluding remarks

By many, the exponential growth in infection numbers is considered as the worst of all possibilities, and it is assumed to occur immediately when the net reproduction number exceeds 1. The first assumption overlooks the fact that the term exponential growth does not represent a complete and adequate characterization, because there is slow and fast exponential growth (e^*α t*^ with a small or large *α*) as there can be slow and fast linear growth (*ż*(*t*)=*α* with a small or large *α*). Accordingly, rapid linear growth can be worse than slow exponential growth over a long period of time. So the growth of *Z*(*t*) in the world was almost linear from the beginning of April to the middle of May, but unfortunately at very high speed. That the second assumption is wrong can be seen from Eqs. (25)-(26). According to them *A*(*t*)=0 for *R*_*z*_(*t*)=1. If *R*_*z*_(*t*) were to become a little larger from this value, then *A*(*t*) would have to jump to 1 immediately for exponential growth, which is impossible according to Eq. (26). ^4^ As we have seen, the modified SIR-model allows for all growth possibilities, from sub-linear up to super-exponential. How dangerous the current growth is depends, however, not only on its type as characterized by *A*(*t*), but also on its current growth rate *v*_*z*_, and on how long this kind of growth will last.

A question, that has been asked many times and the answer to which is important for the acceptance of restrictions that must be endured to contain the pandemic, is: Why do the latter still have to be maintained when the rate of new infections (i.e. *v*_*z*_) has become sufficiently low. The answer to this is not easy, but there is an analogy familiar to all of us which maybe contributes to some understanding. Imagine driving a car and pressing the accelerator halfway to maintain a certain speed. If you want to drive faster, you have to push it further, e.g. three quarters. If you would release the pedal now, you would fall back to lower speed. In a similar way, you either have to maintain the measures undertaken for keeping low the number of new infections, or to replace them with equally effective, but more targeted and less restrictive measures. Note that this analogy is of a more symbolic kind, because neither frictional forces nor inertia and energy supply can be attributed to the pandemic. At most, in the case *R*_*z*_*<*1 or *A*(*t*)*<*0 a constant negative acceleration 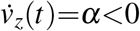 (treated at the end of Section 2.5) can to a limited extent be compared with the acceleration of a car. However, according to Eq. (5), independently of *α* the final velocity *v*_*z*_ is always zero due to the lack of friction. The fact that it is so difficult to bring and keep *R*_*z*_ well below 1 or *A*(*t*) far below zero can possibly be explained as follows: Either the effort to maintain a negative acceleration increases with decreasing *v*_*z*_(*t*), or else, there are processes that work like negative friction. A third mechanism is discussed in the next paragraph.

When looking at the infection rates which the JHU releases for the different countries of the world, one notices that for those who have weathered the crisis rather well (e.g. Germany, Austria, Japan, South Korea or New Zealand), *ż*(*t*) does not decrease completely down to zero, but only to a low and almost constant level. One reason for this could also be, that the different countries do not form closed systems, so that due to the pandemic nature of the covid-19 crisis infections are always brought in from outside. Since the SIR-model only applies to closed systems, this factor cannot be treated with it. That would be possible for the world as a whole, but the latter is still far from low infection rates.

## Data Availability

All data used can be accessed in the Internet.

https://de.wikipedia.org/wiki/SIR-Modell

https://www.arcgis.com/apps/opsdashboard/index.html\#/bda7594740fd40299423467b48e9ecf6

https://interaktiv.tagesspiegel.de/lab/sars-cov-2-das-virus-in-echtzeit/

https://www.welt.de/vermischtes/article206504969/Corona-Wieder-unter-400-neue-Faelle-in-Deutschland-gemeldet.html

## Acknowledgements

For calculations and plots MATHEMATICA 10 was used.

## Footnotes

1 If there are 2 equations in one line, the first is labeled a and the second b.

2 The number of unreported cases can be roughly estimated from the ratio of death cases to the total number of officially registered infected individuals. On July 4, 2020, the apparent death rate caused by covid-19 was 4.7 % for the world and 4.6 % for Germany. According to preliminary informations provided by the virologists, some of them based on statistical surveys but with some uncertainty due to the low number of cases, the true death rate (lethal rate) is only about 0.5 %. In order for this percentage to come out for the populations mentioned, the number of reported cases e.g. in the world must be multiplied by the factor *δ* =4.7*/*0.5=9.4.

3 It suffices to consider the function *a*(*z*) because it runs through the same values as *a*(*t*) and increases monotonically with *t* because of *ż*(*t*) ≤ 0.

4 This does not conflict with the fact that according to Eqs. (58)-(59) *v*_*z*_(*t*) and *z*(*t*) grow exponentially for constant *R*_*z*_*>*1, because also *A≥*1 in this case. The consequence of this is that *R*_*z*_ may not be constant. An example for *A*(*t*)*<*1 with simultaneous *R*_*z*_(*t*)*>*1 is provided by *z*(*t*)=*α t*^2^*/*2, for which *A*(*t*)=1*/*2 and *R*_*z*_(*t*)=(1−1*/t*)^−1^.

## Notes

### Competing Interest Statement

The authors have declared no competing interest.

### Funding Statement

no funding

### Author Declarations

The study is of a theoretical nature with a lot of mathematics so that an evaluation by an ethics committee is irrelevant.

